# Cross-Sectional Analysis of the Association Between Abnormal Immune Responses in the Pathogenesis of Postpartum Cardiomyopathy in Ethiopian Women: A Hospital-based Study at Saint Paul Hospital

**DOI:** 10.1101/2024.09.13.24312550

**Authors:** Azarreia Paulos, Leul Mulu, Kalab Metiku

## Abstract

1.0

**Background:** Postpartum cardiomyopathy (PPCM) is a condition that is characterized by the weakening of heart muscles which is known as left ventricular dysfunction and occurs during the later stages of pregnancy or during the early stages of postpartum.

**Objective:** To evaluate the role of abnormal immune responses and risk factors such as advanced age, high parity, and presence of gestational diabetes in the pathogenesis of PPCM in Ethiopia.

**Method:** A cross-sectional hospital-based study was conducted at Saint Paul Millennium Medical College (SPMCC) between January 2021 and January 2023. All female pregnant patients who were attending the Maternal Care Fetal Unit, Antenatal Unit, and High-Risk Unit at SPMMC were included in this study. However, among those patients, those mothers younger than 20 years old and older than 40 years old were excluded from the study. The total sample size collected from this facility was 250 patients.

**Results:** Oxidative stress and inflammation were primarily correlated with pre-eclampsia and gestational diabetes. These risk factors were particularly prevalent within patients diagnosed with PPCM, 52.8% and 40% respectively. Prolactin Cleavage and Microvascular Dysfunction was associated with higher BMI and advanced age. With a general increasing trend observed in PPCM incidence and advancing age. As well as a high prevalence rate of PPCM in patients with Obesity Class I (at 33.0%). Another notable risk factor was high parity ( 45.0% with 4+ prior pregnancies). The most common symptom presented was dyspnea at 35.0%. The mortality rate was 14% whilst recurrence and the fully recovered ratio were at 13.1% and 34.0%, respectively.

**Conclusion:** Results aligned with those conducted in other African countries (e.g. Zambia and South Africa) however mitigating high mortality as well as low full recovery rate will need to be done by earlier diagnosis, effective treatment, and intervention mechanisms. A different study design will need to be conducted (over a longer time period) to support and make findings more holistic.

## 2.0 Introduction

### 2.1 Background

Postpartum Cardiomyopathy (PPCM) is a condition that is caused by the weakening of the heart muscles, leading to heart failure. Furthermore, PPCM is a type of congestive heart failure that occurs during the later stages of pregnancy or within the first couple of months after the postpartum period (Dugdale and Conaway). This condition’s genesis is still unknown mainly because it is characterized by an abrupt onset in individuals who have never had a heart condition before. Thus there is no identifiable cause of cardiac failure in these women that are diagnosed with PPCM (Dugdale and Conaway).

PPCM is directly correlated with left ventricular systolic dysfunction, which causes weakening and enlargement of the left ventricular heart muscles (Liu). As a result, this compromises the heart’s ability to effectively pump blood to the rest of the body (“Peripartum Cardiomyopathy”). Symptoms vary but remain similar to other forms of congestive heart failure (CHF) including but not limited to dyspnea, fatigue and edema.

Additionally, PPCM and other types of CHF are associated with similar risk factors which include advanced maternal age, high parity (71% of women diagnosed with PPCM had three or more prior pregnancies), high gravidity, twin pregnancy, use of tocolytic therapy (greater than 4 weeks can cause silent ischemia), and hypertension (“Postpartum Cardiomyopathy - StatPearls”).

Studies have shown that PPCM is more prevalent among mothers with African descent. This condition disproportionately affects this demographic due to a variety of factors which is the late diagnosis, poorer systolic function on diagnosis with larger cardiac dimensions, and less favorable outcomes (Strasserking). At 6-month follow-up, only 37 % of African patients had recovered compared to 57 % and 62 % in Europe and Asia-Pacific respectively. This supports other data that showed women of African descent do not fare as well as their non-African counterparts(Strasserking).

Saying this, the abnormal responses that predispose mothers to PPCM could be attributed to a few biological processes. These processes are specific forms of abnormal response called Oxidative stress, Inflammation, Prolactin Cleavage and Microvascular dysfunction

#### Oxidative Stress and Inflammation

Concerning abnormal immune response, a mother goes through a variety of hemodynamic and hormonal changes during all trimesters of pregnancy (e.g. increased cardiac output, and decreased blood pressure). These molecular changes trigger a range of immune responses some of which could predispose a mother to further complications.

In PPCM, oxidative stress and inflammation are two biological processes that are intertwined and play a significant factor in the pathogenesis of pregnancy complications(Scarian). These systems are important in cellular integrity and defense against pathogens. Oxidative stress occurs when there is a dysfunctionality or a disruption in the equilibrium between the generation of reactive oxygen species (ROS) and the body’s ability to neutralize the ROS’ that are created (Scarian). ROS plays a crucial role in cell proliferation as well as protection against pathogens (Cohen). However, unmonitored and excessive generation of these species results in damage to various subcellular components (Lipids, proteins, and DNA) which long-term leads to oxidative stress and the pathogenesis of chronic conditions (Dugdale and Conaway).

Inflammation is a defense mechanism where the body responds to harmful stimuli such as pathogens. It is a systemic reaction by the body in effort to identify, remove, and heal harmful agents with respect to cell tissue damage (Chen). It could be classified as both acute and chronic inflammation. Acute inflammation is characterized by redness, swelling, heat, pain, and loss of function that occurs around the affected area. In contrast, chronic inflammation occurs over a longer period of time inherently leading to tissue damage, specifically within the heart. This leads to diastolic functionality in the left ventricle which comprises the heart’s ability to pump sufficient blood, which is also known as PPCM (Chen).

#### Prolactin Cleavage and Microvascular Dysfunction

Another factor that measured immune responses was prolactin Cleavage and microvascular dysfunction. Prolactin (PRL) is a hormone that has a variety of different roles. Specifically within the endocrine and immune systems (Zandman et al.) Its responsibility primarily lies within lactation, but it can also act as a cytokine, impacting immune responses. PRL affects the immune system at the levels of innate and adaptive immunity. During pregnancy, under specific conditions, through the guidance of a specific enzyme (cathepsin D), Proclcatin cleavage occurs resulting in the formation of 16-kDA fragment (Zandman et al.). These have specific properties that increase cell death while inhibiting the production of new blood levels. As such this prolactin cleavage results in myocardial injury and remodeling, which is relevant in the context of the pathogenesis of PPCM.

Microvascular dysfunction is another crucial role player in the prevalence of PPCM. This condition is linked to chronic inflammation and oxidative stress, factors that cause endothelial dysfunction. This hinders vasodilation and increases vascular permeability, which resultantly halts regular microvascular function (Zandman et al.). Additionally, microvascular dysfunction causes the synthesis of fibroblasts, which hence results in collagen deposition and fibrosis within the myocardium, hindering the heart muscle’s contractile function and contributing to the development of PPCM (Scarian)

This research aims to investigate, analyze, and deduce the primary abnormal immune response that occurs during pregnancy and how exactly these conditions influence the prevalence of PPCM within a hospital setting.

### 2.2 Statement of the problem

PPCM remains to be increasingly prevalent worldwide with rates on a steady rise (citation). In Nigeria for example, it is reported that there has been a notable increase in numbers with the incidence being at about 1 out of 100 life births (Riyami). Additionally, a usually undermined, yet very critical aspect is adequate healthcare care access which influences maternal health. This causes delays such as diagnostic period, intervention, and management strategies thus escalating such conditions overall and giving rise to mortality rates.

Ethiopia’s high maternal mortality ratio and status as the second most populous country on the African continent are serious drawbacks for maternal health. The number of maternal fatalities in a specific time period per 100,000 live births over the same time period is known as the maternal mortality ratio, or MMR (“Maternal mortality ratio (per 100 000 live births)”). Ethiopia’s MMR remained high, declining from 708 per 100,000 live births in 1990 to 497 per 100,000 live births in 2013, despite the country’s population growth (Tessema). The top five causes of death for mothers in 2013 were determined to be PPCM, which accounted for 25.7 percent of deaths; complications from abortions, which were responsible for 19.6% of deaths; maternal hemorrhage, which was 12.2% of deaths; hypertensive disorders, which comprised for 10.3% of deaths; and infections, which included hepatitis, malaria, influenza, and tuberculosis and killed 9.6% of the women (Tessema). The majority of maternal fatalities occurred in the postpartum phase and in the age range of 20 to 29 (Tessema).

According to a study utilizing data from the 2016 Ethiopian Demographic and Health Survey (EDHS), 62% of expectant mothers attended at least their first prenatal visit; 28% of women received skilled birth attendant assistance during childbirth; 17% received postnatal care; and 35%, depending on the region, used a contraceptive method (Tessema). The MMR is still high and maternal health services are still insufficient, indicating that additional coordinated efforts are needed to improve the likelihood of safe motherhood, particularly in the postnatal period. This remains to be a primary driver of MMR and the overall prevalence of cardiac issues within Ethiopia as well as other Low and Low-to-Middle Income countries (LIC and LMICs).

### 2.3 Significance of the study

PPCM is a condition with an unknown pathophysiology. Thus whilst the prevalence of this condition seems to be on the rise, the etiology and cause remain unclear. The lack of adequate diagnostic facilities and awareness about the disease contribute to the rising number of patients with PPCM within the Ethiopian demographic. This study will be helpful in filling the gap of information on the magnitude, pattern, and associated factors of PPCM among pregnant mothers with respect to abnormal immune response at Saint Paul Hospital in Addis Ababa governmental hospitals. It might be important to know the magnitude and patterns of PPCM at Addis Ababa governmental hospitals to develop strategies for improved patient management and rehabilitation. This may help patients to get early treatment and appropriate management. This may include medical and surgical interventions as well as providing adequate counseling to mothers and their respective families. Information on associated factors may shed light on their roles as risk factors for the occurrence of PPCM hence it provides baseline data for future detailed studies and public health strategic plans.

## 3.0 Methodology

### 3.1 Study Design and Biomarkers

This study was an unmatched hospital-based cross-sectional study design that was conducted at St Paul and was conducted in St Paul Medical Millennium College (SPMMC). Specifically within the high-risk pregnancy units (HRUs) and high-risk antenatal units (AU) and Maternal and Child Fetal Care Units (MCFU). This facility provides prenatal care, antenatal check-ups, labor and delivery management, postnatal care, and neonatal intensive care. As well as diagnosis and early detection of anomalies in the health of pregnant mothers and post-natal.This study was conducted from January 2021 to January 2023. Currently, this facility has 700+ beds, with an annual average of 200,000 patients and a catchment population of more than 5 million thus being one of the biggest referral hospitals in the region.

Biomarkers that were measured in this study in relation to oxidative stress and inflammation were blood pressure ranges (looking at the presence of high blood pressure i.e Hypertension Stage 1/2) Blood pressure ranges and the absence or presence of gestational diabetes during pregnancy. Additionally, when analyzing the effect of prolactin cleavage and microvascular dysfunction on the prevalence of PPCM, Parity as well as BMI values (Obesity) were used as biomarkers.

### 3.2 Study populations (Inclusion and Exclusion criteria)

All female pregnant patients who were attending the MCFU, AU, and HRUs at PSMMC were included in this study. However among those patients, those mothers ≤ 20 *y/o or* ≥ 40 *y*/*o* as were excluded from the study. The total sample size collected from this facility was 250 patients.

### 3.3 Ethical Approval

Ethical clearance was obtained from the Institutional Review Board of The International Community School of Addis Ababa; Permission was also obtained from each facility for doing the study. Additionally, written consent was obtained from each mother after providing sufficient information on the purpose of the study; and parental written informed consent was obtained from the family/caregivers. Confidentiality was assured for all the information provided and no personal identifiers (anonymity) will be used on the questionnaires. It is believed that there is no anticipated harm for the clients except for their time scarification at the time of data collection. The collected data will be kept in a secure place until the publication of the result.

### 3.4 Data collection procedure

In order to collect data on postpartum cardiac anomalies in mothers between 20 and 40 y/o the structured checklist was administered. The checklist was first developed in English and translated into Amharic (the Ethiopian official language), and then translated back into English by a third person to check the reliability of the checklist. Data collectors were trained for two days, regarding the objectives of the study, and on sampling procedures. Data on socio-demographic and clinical information was gathered from the respective patients’ cards. The diagnosis of PPCM of the mother was taken from the children’s medical records after it was confirmed by a cardiologist with an echocardiograph. During data collection Cardiologists, Gynecologists, and Perinatologists were consulted when there was unclear diagnosis. Moreover, a diagnosis was excluded if it wasn’t confirmed by Perinatologist specialists.

Additionally, a questionnaire list was created and implemented in the patients included in the study. The questionnaire list was designed to gather relevant information on the patient’s prenatal health conditions concerning hemoglobin levels, any previous pregnancies, or other health complications that occurred before the specific pregnancy term.

## 4.0 Results

**Table 1:** Data of Patient at Saint Paul Hospital (Presented as n)

| Age Ranges, n (%) |  |
| --- | --- |
| 20-25 | 25 (10.0%) |
| 26-30 | 95 (38.0%) |
| 31-35 | 78 (31.0%) |
| 36-40 | 52 (21.0%) |

| Parity, n (%) |  |
| --- | --- |
| 1 | 62 (25.0%) |
| 2-3 | 75 (30.0%) |
| 4+ | 113 (45.0%) |

*Incidence of PPCM occurred mostly within the 26-30 age group (38%), followed by ages of 31-35 (31%) then 36-40 (21%), and then lastly 20-25 (10%). Additionally, 113 out of 250 mothers reported having 4+ total pregnancies. 30% of mothers (n=75) reported 2-3 total pregnancies and the incidence of PPCM was least prevalent among those mothers who have only been pregnant once (25%) .*

**Table 2:**
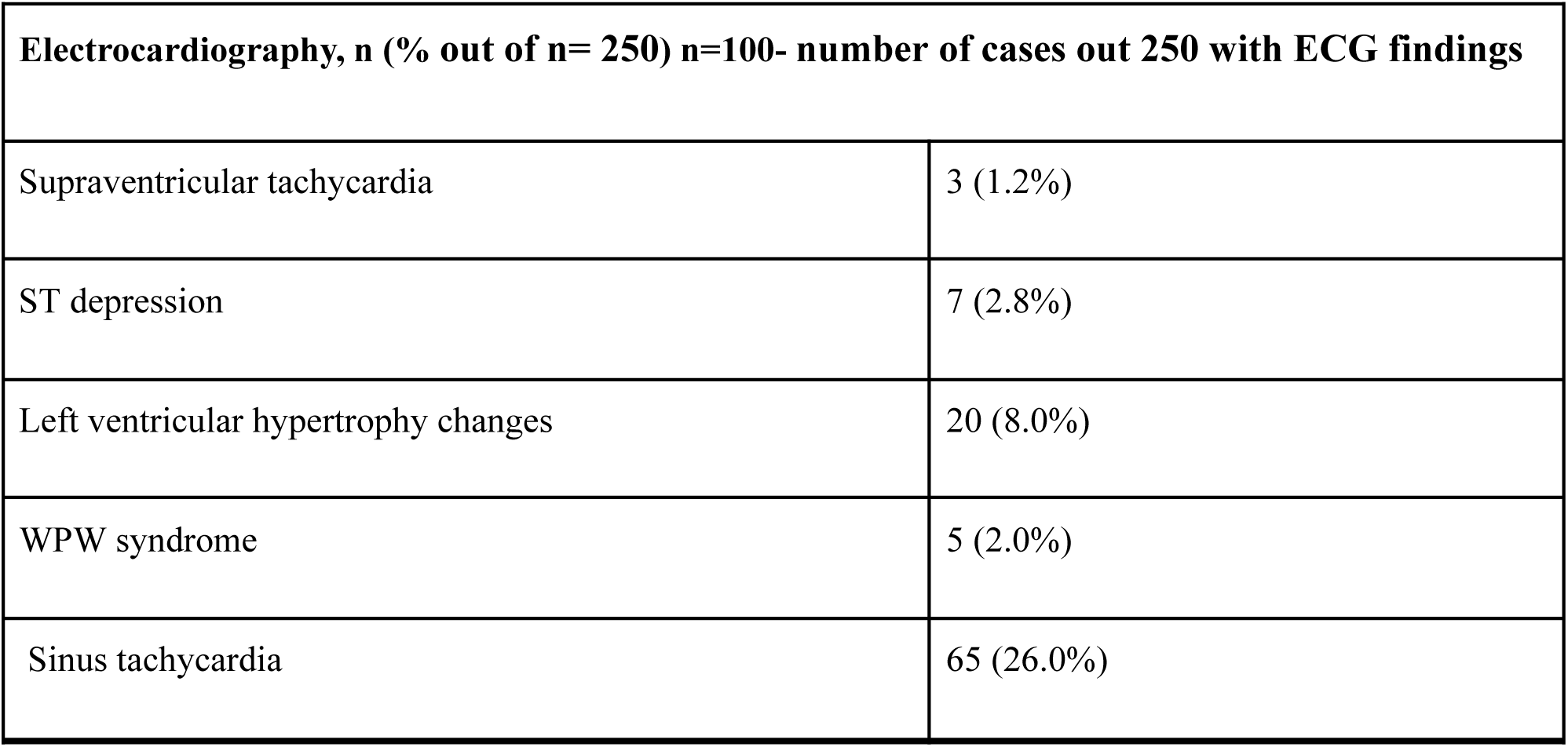
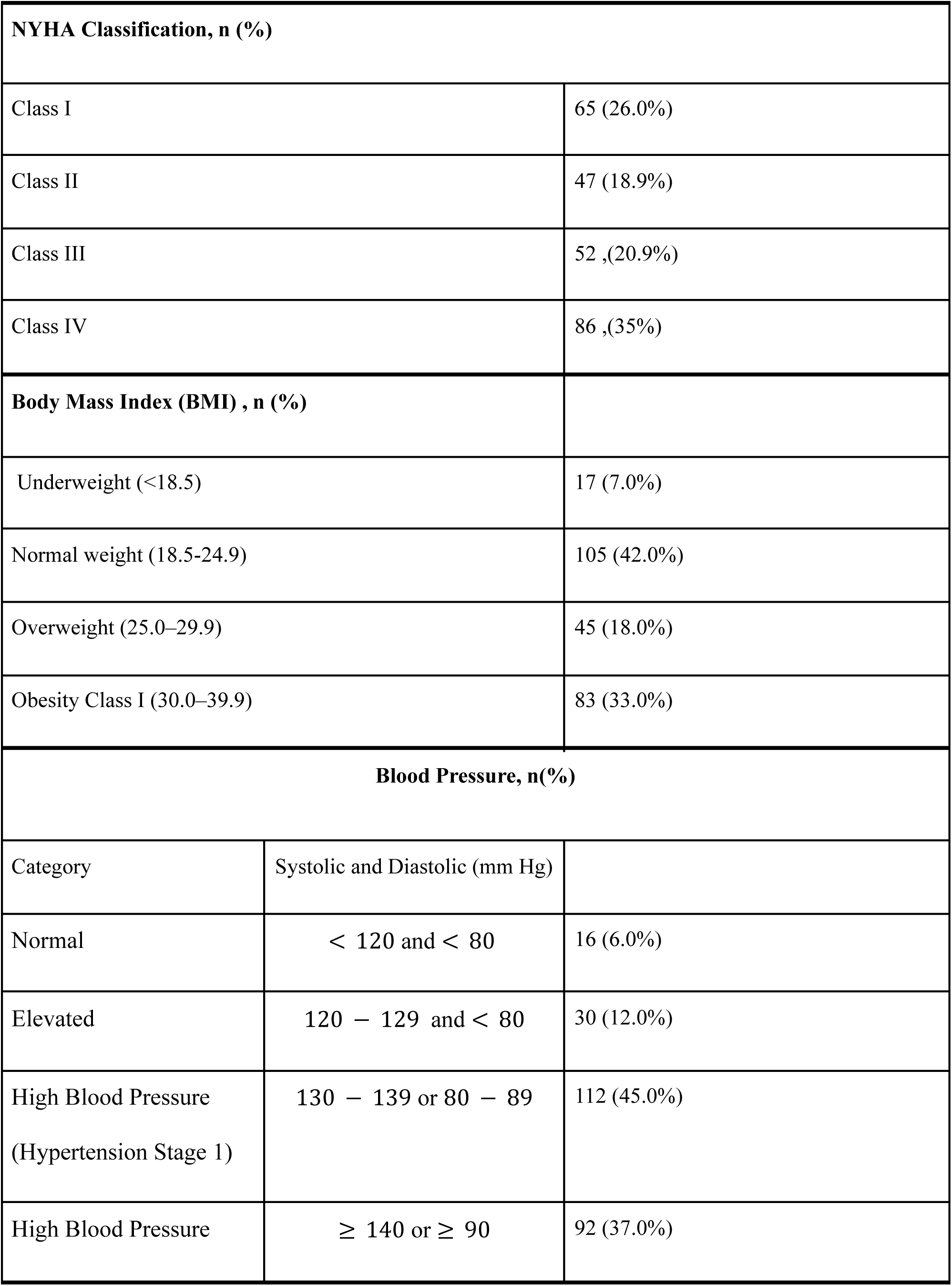
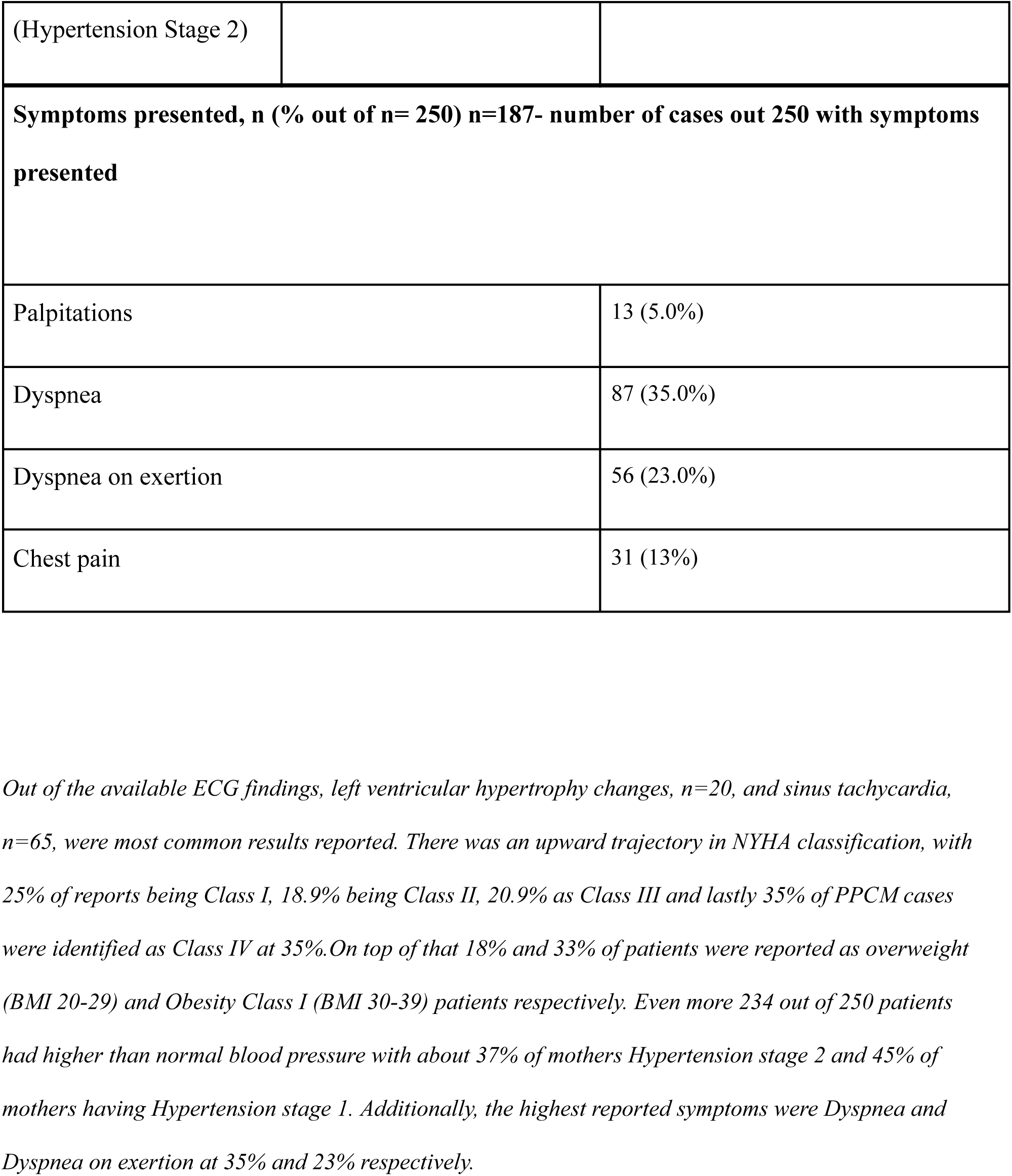
Clinical Characteristics of Patients at Saint Paul Hospital (presented as n)

**Table 3:** Antenatal Comorbidities of Patients at Saint Paul Hospital (Presented as number of patients, n)

|  |  |
| --- | --- |
| **Prior Cardiac related issues, n (%)** |  |
| Yes | 162 (65%) |
| No | 88 (35%) |
| **Hypertensive disorders during pregnancy, n(% out of n= 250) n=204- number of cases out of 250 with hypertensive disorder diagnosis** |  |
| Pre-eclampsia | 132 (53%) |
| Chronic Hypertension | 72 (29%) |
| **Diabetes in pregnancy, n (%) n=100** |  |
| Gestational diabetes | 66 (22.0%) |
| Pre-existing diabetes | 34 (14%) |

*The majority of mothers were found to have prior cardiac-related issues (n=162). Of the 204 mothers who had hypertension, 132 patients were diagnosed with preeclampsia while 72 mothers had Chronic hypertension. The majority of mothers with diabetes had gestational diabetes n=66 whilst the majority of them had pre-existing diabetes n=34.*

**Table 4:** Perinatal Outcomes of Patients at Saint Paul Hospital.

|  |  |
| --- | --- |
| **Admittance to ICU, n (%)** | 24 (10%) |
| **Fully recovered , n(%)** | 85 (34.0%) |
| **Recurrence, n (%)** | 33 (13.%) |
| **Mortality, n(%)** | 35 (14.0%) |

*Mortality was at 14% whilst 85 out of 250 patients fully recovered. 13.0% recurrence rate and 10% of mothers were admitted into ICU whilst 33 mothers had recurrence of PPCM.*

**Graph 1.1.**
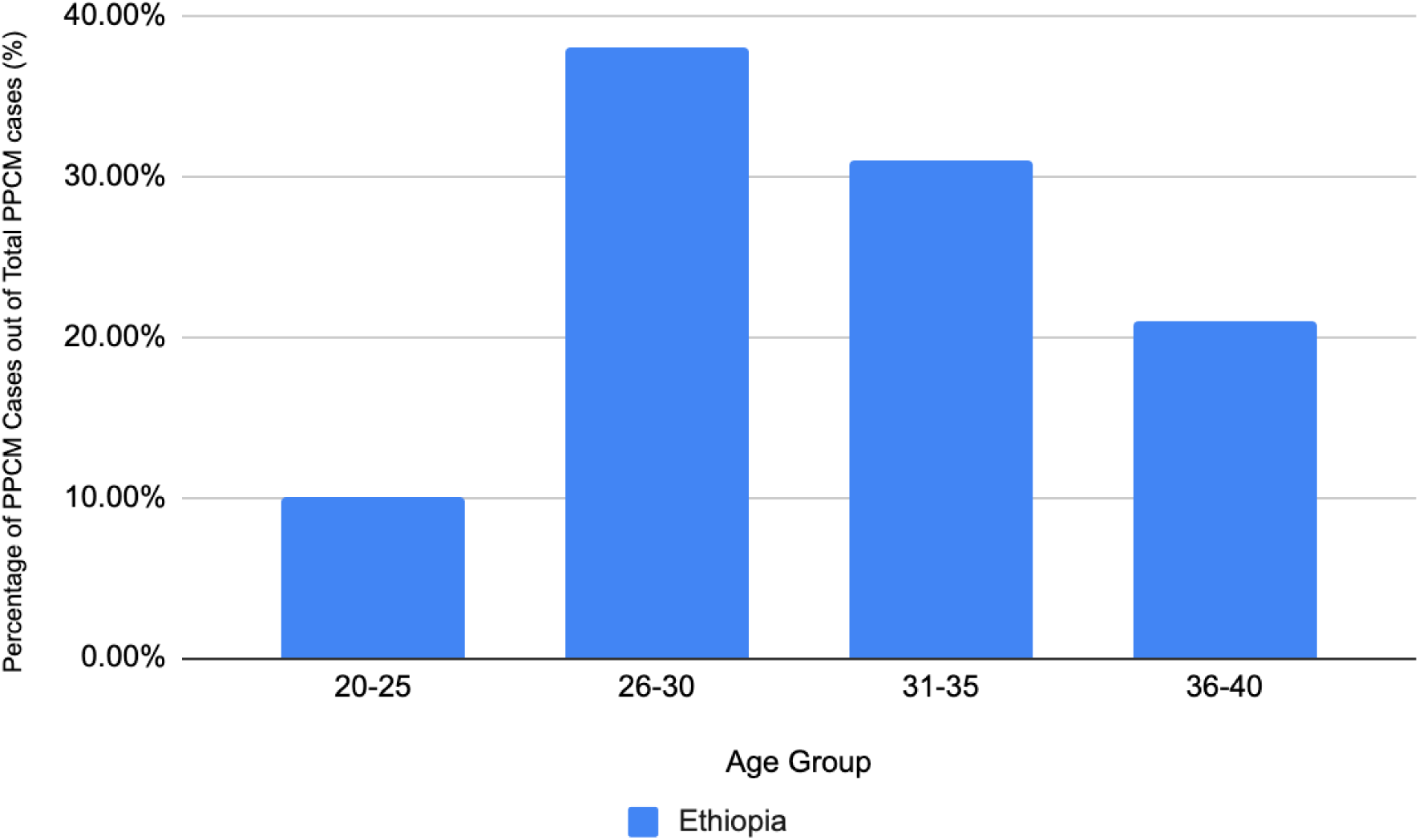
Prevalence of CHF within each age group within Saint Paul Hospital

**Graph 1.2.**
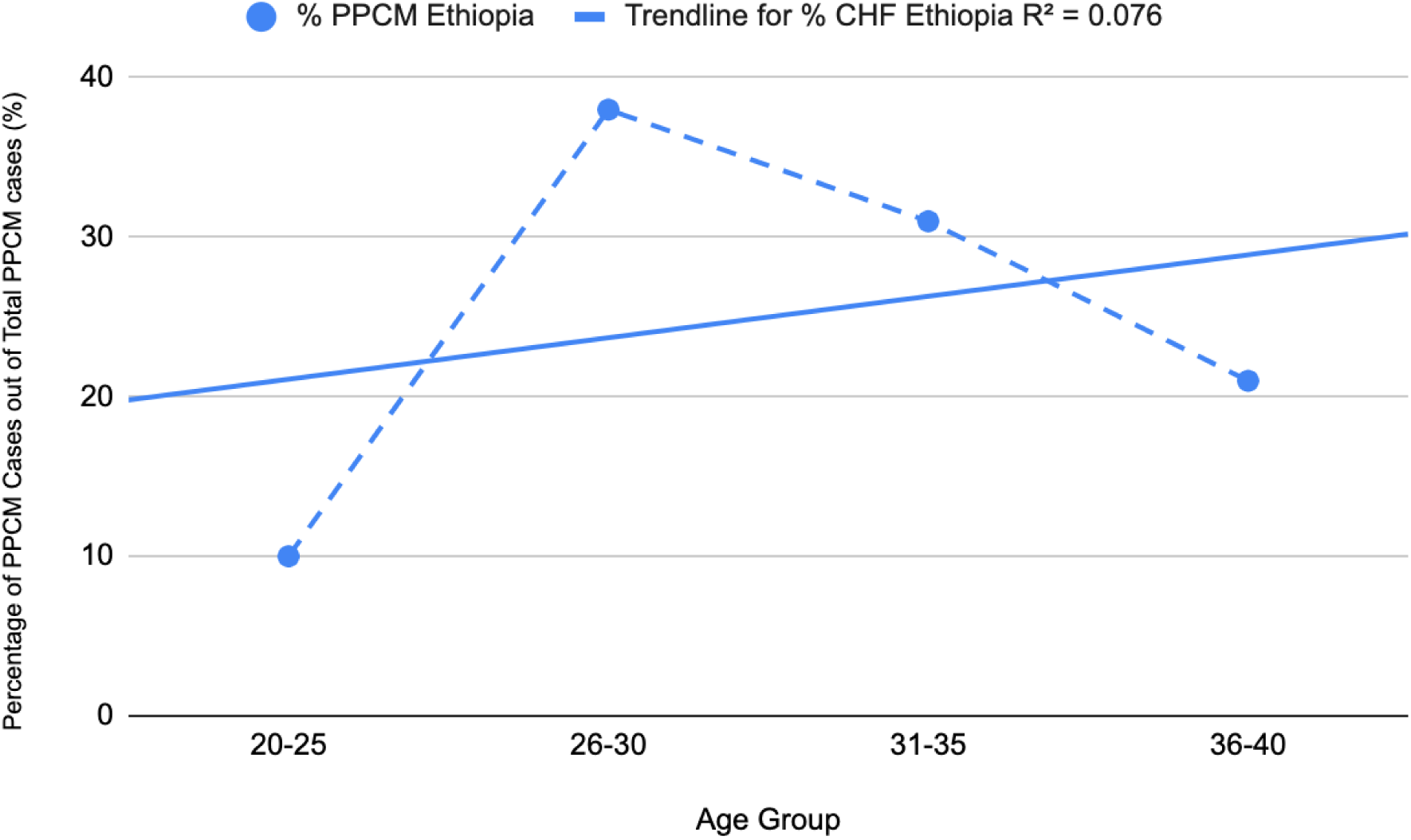
Prevalence of PPCM within each age group along with *R*^2^ *value* within Saint Paul Hospital

**Figure 1.1.**
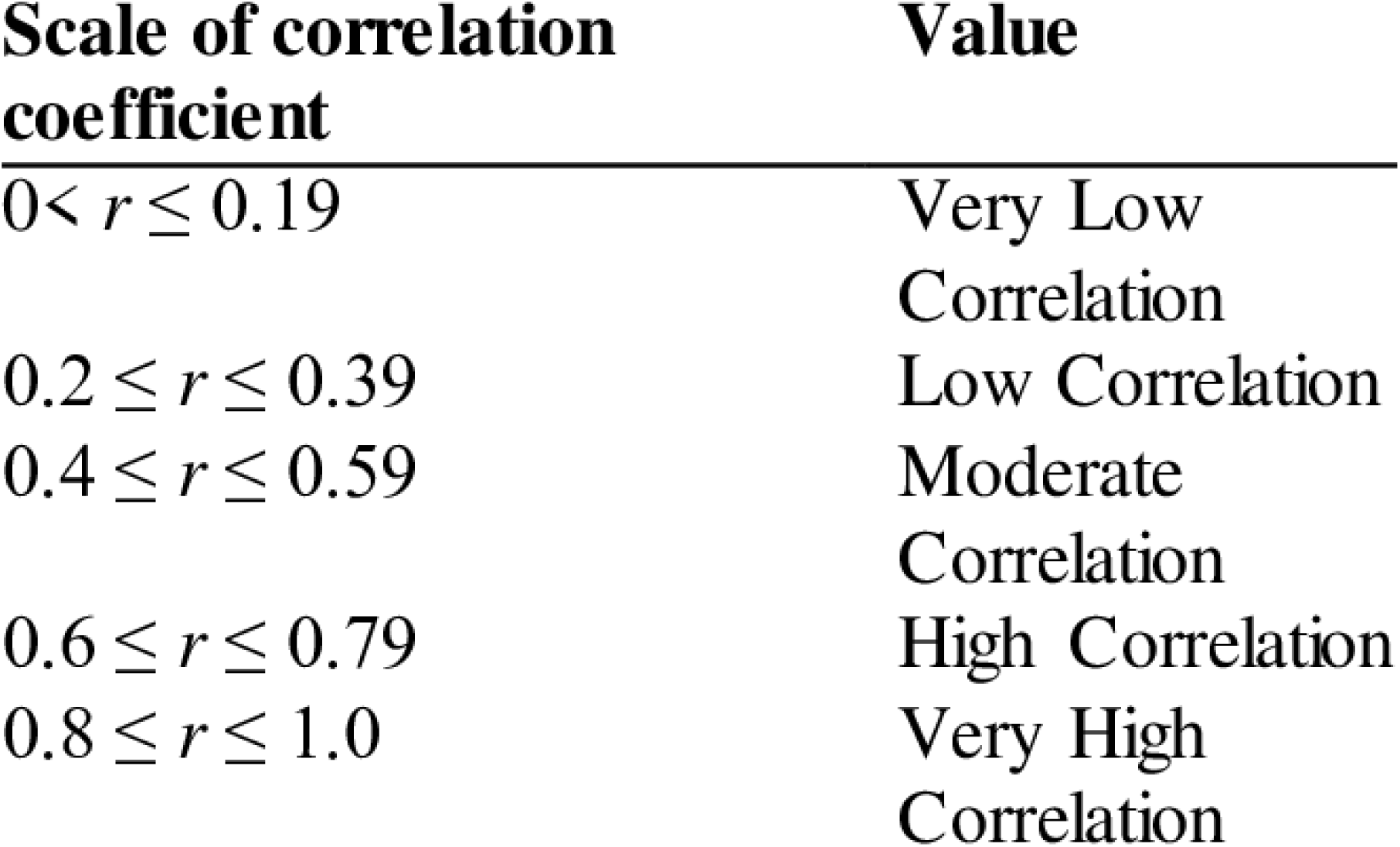
The scale of Pearson’s Correlation Coefficient and correlation determination

### Statistical Analysis

A Pearson’s correlation coefficient test was conducted. This was done in order to measure the correlation (if any) between Age and incidence of PPCM within Saint Paul Hospital. Meaning how would a change in age (x) affect the prevalence of PPCM (y). Results provided a *R*^2^ = *0.07* (As seen in Graph 1.1). This means that there is a very low correlation between x and y. To be more exact, a change or increase in x didn’t affect a change in y. After close consideration nevertheless, this could be primarily due to an outlier in the data (Age group 26-30) because the data trends in general demonstrate a correlation between the two (observed by the linear trend line). However, it could be determined from data in Graph 1.2 that the exact age group that is more susceptible to the incidence of PPCM is 26-30, followed by 31-35 then 36-40, and lastly 20-25.

## 5.0 Discussion

### Oxidative Stress and Inflammation

Oxidative stress and inflammation go hand in hand and are known to have what is called a bi-directional relationship. This means that oxidative stress can cause inflammation through the generation of redox-sensitive pathways which leads to the synthesis of pro-inflammatory cytokines(Scarian). Similarly, Inflammation can cause oxidative stress. This is done through the production of ROS that exacerbates oxidative stress. As such this demonstrates their bidirectional nature and in fact how they play a combined role in the pathogenesis of abnormal immune responses and eventually, the development of PPCM.

There are a few factors that influence oxidative stress as well as inflammation stress levels. These are Blood pressure ranges, and the absence or presence of gestational diabetes during pregnancy (Renuga Sundari).

Elevated blood pressure, specifically when classified as hypertension (where systolic and diastolic is ≥ 130-139 or 80-89 mm Hg as seen in *Table 2)* is directly correlated with heightened oxidative stress and inflammation within the body(Gumilar). This is because the increase in blood pressure is associated with endothelial dysfunction (Gumilar). A condition that is characterized by the dysfunction of the inner lining of blood vessels as they become less responsive to a variety of chemical signals for dilation and construction (Gumilar). Resulting in vascular resistance, which hence further increases blood pressure. On top of that, there is also a link between endothelial dysfunction, systemic inflammation, and oxidative stress. This is because damaged endothelial cells synthesize pro-inflammatory cytokines and ROS. This results in unnecessary stress and pressure on the cardiovascular system and damage to heart tissue and muscles which would eventually result in the pathogenesis of PPCM (Gumilar).

Keeping this in mind, when observing data from patients in Saint Paul Hospital (SPH) in *Table 2*, it can be observed that there is a direct correlation between blood pressure level and prevalence of PPCM. This means an increase in blood pressure would also cause an increase in the incidence of PPCM. Although of those patients diagnosed with PPCM, most of them reported a blood pressure level between 130-139 or 80-89 mm Hg (45%), a general trend of this positive correlation can be observed. The incidence among patients with “Normal” and “Elevated blood pressure” was reported to be relatively low, with values at 6% and 12% respectively. Conversely, 37% of patients with PPCM were reported to have high blood pressure specifically (hypertension stage 2). Additionally, of the 204 patients who reported either hypertension stage 1 or hypertension stage 2, 132 were found to be pregnancy-induced hypertension. Suggesting that from the entire patient population with PPCM diagnosis 52.8% were found to be pre-eclampsia. Pre-eclampsia is a condition referred to as “pregnancy-induced” hypertension after 20 weeks of gestation and is also associated with endothelial dysfunction and systemic inflation (Dugdale and Conaway). This shows how high blood pressure increases susceptibility to PPCM and is in fact a primary risk factor in the pathogenesis of PPCM.

Furthermore, another factor closely linked to oxidative stress and inflammation is the presence of gestational diabetes GDM. Gestational diabetes is diabetes that arises during pregnancy. Specifically through raised insulin levels and hyperglycemia. This results in insulin resistance where the body’s cells become unresponsive to insulin. (Xavier). The inability of pancreatic β-cells to compensate adequately for this insulin resistance results in the development of GDM. Hyperglycemia exacerbates oxidative stress through the over generation of reactive oxygen species which results in higher inflammation levels (Xavier). GDM is linked to PPCM in a variety of ways: Endothelial Dysfunction, Myocardial damage, and impaired immune regulation (Xavier). All through this the ability of the heart to pump out sufficient blood to the body is compromised, presenting itself as a significant risk factor in the onset of PPCM.

Similar trends can be observed in patient data from SPH, with a staggering 40% of patients with PPCM also being diagnosed with either pre-existing or gestational diabetes. Additionally, there is a clear link between Gestational diabetes and PPCM as 22% of patients reported cases of GDM in comparison to 13.6% of pre-existing diabetes. Highlighting the presence of GDM and hypertension as factors that influence the prevalence of PPCM

### Prolactin Cleavage and Microvascular Dysfunction

Aging is associated with increased oxidative stress and inflammation as well as increased endothelial dysfunction. As such with advanced age there is a general susceptibility to PPCM (“Postpartum Cardiomyopathy - StatPearls”). This seems to be the general observation within the data at SPH, with the overall increase in PPCM onset among older patients, bearing the significant outlier. This data also corresponds with another study conducted in Oman which examined the *“Incidence, Risk Factors, Maternal and Neonatal Outcomes of Peripartum Cardiomyopathy”,* which saw the highest PPCM cases within a similar age group (middle reproductive age group).

Noting that obesity is a significant risk factor of PPCM, patients with higher BMI values were found to have PPCM with the reported cases being 18% and 33% for classifications of overweight and obesity class 1 respectively. Additionally out of reported 250 cases, 45.0% of cases were of mothers that had 4 or more prior pregnancies. Corresponding with existing data on high parity as a risk factor in relation to the pathogenesis of PPCM.

In correspondence with the New York Heart Association Classification (NYHA Classification), 86 (35.0%) of patients were categorized as Class IV, then Class I in 65 (26.0%), followed by Class III in 53 (20.9%), and finally Class II in 47 (18.9%). Notably high NYHA classification in class IV could be attributable to late diagnosis due to limited health care facilities. Additionally, the majority of patients traveled from rural Ethiopia to Addis to receive medical attention as a result potentially delaying diagnosis. Next, only 100 patients out of 250 had taken ECG records. Even after follow-up, limited ECG machines meant that availability for such scans was limited. However, of those recorded, Sinus tachycardia was found to be the most common reported at 56 cases.

With respect to symptoms presented, 187 out of 250 cases were ones with symptoms. Most common symptom presented was Dyspnea with 87 out of 187 cases. Findings were consistent with a study conducted in Zambia with 44 (the highest number of symptoms reported), which evaluated “*Characteristics and outcomes among women seen at a referral hospital in Lusaka, Zambia”* (Strasserking).

There was a relatively low percentage of individuals within this study that fully recovered 34%. This was in correspondence with a 6-month follow-up cohort study conducted in South Africa, of which 32% of patients fully recovered (Azibani). Nevertheless, this low rate is concentrated by similar studies conducted in Oman and Germany, with both studies presenting 56% and 52% respectively (Azibani). Similarly, in this study, there was a relatively high mortality of 23.0% which was similar to that of the study conducted in South Africa (11%). And is significantly higher than studies done in Oman (3.4%).

PPCM is a multi factored condition with a wide range of different risk factors. However among these conditions there are foundational principles that seem to be similar. High blood pressure and Diabetes are conditions that predispose mothers to PPCM as well as a variety of cardiovascular diseases. Lifestyle and dietary changes among women, specifically mothers is one way to which the incidence of PPCM can be mitigated. According to recent studies, a sedentary lifestyle as well as a poorly managed diet is a factor of rising blood pressure as well as presence of diabetes (Sami). As such this has an effect on PPCM incidence. Sedentary lifestyle has been on a rise in Ethiopia, this has also been similar with global trends (Motuma).

There are some of the ways whereby the risk factors of PPCM can be managed with the view of minimizing the incidence of PPCM. Adopting necessary dietary and lifestyle changes would help reduce incidence of PPCM risk factors. In addition to this, this approach does not only address immediate health issues, but it also lays a foundation for cardiovascular health wellness among the Ethiopian population.

Additional studies will need to be conducted in order to measure the effectiveness of changing lifestyle and dietary habits in relation to incidence of PPCM.

## 6.0 Conclusion and Evaluation

### Conclusion

Postpartum Cardiomyopathy seems to be on the rise in East Africa with rising prevalence. This study identified, evaluated, and analyzed the role of gestational diabetes, high parity, blood pressure levels, BMI as well as pre-eclampsia in the incidence of PPCM in relation to abnormal immune responses. There were many similarities with other studies conducted within African countries and countries of similar socioeconomic status in comparison to Ethiopia however when observed in international studies Ethiopia is unfavorable in terms of relatively high mortality, and low recovery rates of PPCM.

To be exact, from the study population, 22% of mothers were reported with gestational diabetes. Whilst a high 52.8% patients had pre-eclampsia (pregnancy-induced hypertension).113 out of 250 mothers had had 4+ pregnancies and 51% of PPCM cases mothers BMI indicated either Obesity Class I or Overweight.

Further studies are necessary to effectively evaluate and close knowledge gaps revolving around the etiology of PPCM.

### Evaluation

#### Strengths

Conducting both prospective and retrospective study allowed for improved accuracy and recency of patient data. Had the study purely relied on retrospective data, inaccuracies in card registration and missing clinical characteristics would have not been compensated for.

The facility selected was a referral hospital as such there were patients from all over Ethiopia. This meant that out of the sample size patients were from all over Ethiopia. As a result, depicting a fair representation of PPCM incidence within Ethiopia as a whole.

To my knowledge, this is the first paper that evaluates PPCM within Ethiopia. Specifically evaluating abnormal immune response and other risk factors in the pathogenesis of PPCM.

#### Limitations

The chosen study design (cross-sectional) meant data was limited by time period. As such this could’ve resulted in misrepresentation of values had time period been not a limiting factor. Thus analysis of PPCM through another study design would holistically represent findings.

## Data Availability

All data produced in the present study are available upon reasonable request to the authors.

## Notes

### Competing Interest Statement

The authors have declared no competing interest.

### Funding Statement

This study did no receive any funding

### Author Declarations

Institutional Review Board of the International Community School of Addis Ababa as well as Saint Paul Medical Millennium College gave ethical approval for this work.

## References

Azibani, Feriel. “Outcome in German and South African peripartum cardiomyopathy cohorts associates with medical therapy and fibrosis markers.” Wiley Online Library, https://onlinelibrary.wiley.com/doi/10.1002/ehf2.12553.

“Burden, predictors and short-term outcomes of peripartum cardiomyopathy in a black African cohort.” NCBI, 21 October 2020, https://www.ncbi.nlm.nih.gov/pmc/articles/PMC7577461/. Accessed 7 July 2024.

Chen, Linlin. “Inflammatory responses and inflammation-associated diseases in organs.” NCBI, https://www.ncbi.nlm.nih.gov/pmc/articles/PMC5805548/. Accessed 6 July 2024.

Cohen, Alan A. “Beneficial and Detrimental Effects of Reactive Oxygen Species on Lifespan: A Comprehensive Review of Comparative and Experimental Studies.” NCBI, 11 February 2021, https://www.ncbi.nlm.nih.gov/pmc/articles/PMC7905231/. Accessed 5 July 2024.

Demkow, Marcin. “Biomarkers in Peripartum Cardiomyopathy—What We Know and What Is Still to Be Found.” NCBI, https://www.ncbi.nlm.nih.gov/pmc/articles/PMC10813209/. Accessed 6 July 2024.

Dugdale, David C., and Brenda Conaway. “Peripartum Cardiomyopathy - Symptoms and Causes.” Penn Medicine, 5 8 2022, https://www.pennmedicine.org/for-patients-and-visitors/patient-information/conditions-treated-a-to-z/peripartum-cardiomyopathy. Accessed 18 June 2024.

Gumilar, Khanisyah Erza. “Connecting the Dots: Exploring the Interplay Between Preeclampsia and Peripartum Cardiomyopathy.” NIH, https://www.ncbi.nlm.nih.gov/pmc/articles/PMC11219213/.

Karaye, Kamilu M. “Epidemiology of peripartum cardiomyopathy in Africa Authors.” Global Cardiology, https://www.globalcardiology.info/site/article/view/31.

“Peripartum Cardiomyopathy.” American Heart Association, 28 May 2024, https://www.heart.org/en/health-topics/cardiomyopathy/what-is-cardiomyopathy-in-adults/peripartum-cardiomyopathy-ppcm. Accessed 30 June 2024.

“Postpartum Cardiomyopathy - StatPearls.” NCBI, https://www.ncbi.nlm.nih.gov/books/NBK534770/. Accessed 18 June 2024.

Renugasundari, Manoharan. “Inflammation and decreased cardiovascular modulation are linked to stress and depression at 36th week of pregnancy in gestational diabetes mellitus.” NCBI, 26 June 2023, https://www.ncbi.nlm.nih.gov/pmc/articles/PMC10293198/. Accessed 6 July 2024.

Riyami, Nihal Al. “Incidence, Risk Factors, Maternal and Neonatal Outcomes of Peripartum Cardiomyopathy (PPCM) in Oman - Global Heart.” Global Heart Journal, 2 May 2023, https://globalheartjournal.com/articles/10.5334/gh.1198. Accessed 30 June 2024.

Scarian, Eveljn. “New Insights into Oxidative Stress and Inflammatory Response in Neurodegenerative Diseases.” MDPI, https://www.mdpi.com/1422-0067/25/5/2698. Accessed 5 July 2024.

Strasserking, Fiona E. “Peripartum cardiomyopathy: Characteristics and outcomes among women seen at a referral hospital in Lusaka, Zambia.” NCBI, 22 August 2022, https://www.ncbi.nlm.nih.gov/pmc/articles/PMC9421395/. Accessed 18 June 2024.

Tessema, Gizachew Assefa. “Trends and causes of maternal mortality in Ethiopia during 1990–2013: findings from the Global Burden of Diseases study 2013 - BMC Public Health.” BMC Public Health, 2 February 2017, https://bmcpublichealth.biomedcentral.com/articles/10.1186/s12889-017-4071-8. Accessed 30 June 2024.

Xavier, Jadriane Almeida. “Gestational Diabetes Mellitus: The Crosslink among Inflammation, Nitroxidative Stress, Intestinal Microbiota and Alternative Therapies.” NCBI, 7 January 2022, https://www.ncbi.nlm.nih.gov/pmc/articles/PMC8773111/. Accessed 6 July 2024.

Zandman, Gisele, et al. “Prolactin and Autoimmunity.” Frontiers, https://www.frontiersin.org/journals/immunology/articles/10.3389/fimmu.2018.00073/full. Accessed 6 July 2024.

